# Transfer learning for Covid-19 detection in medical images

**DOI:** 10.1101/2022.07.25.22278017

**Authors:** Maryam El Azhari

## Abstract

As of late, the Covid infection 2019 (COVID-19) has caused a pandemic sickness in more than 200 nations, therefore impacting billions of people. To control the spread of the coronavirus, it is crucial to detect infected individuals and ensure their complete isolation to prevent further infection. Chest X-rays and CT-scans have been proven to be very promising as signals of the infection can be clearly shown in lung areas. Transfer learning from ImageNet dataset has become the latent trend in medical imaging applications. However, there are major differences between ImageNet and medical imaging datasets. Therefore, the feasibility of transfer learning in medical applications remains questionable. This paper investigates the performance of five fine-tuned pre-trained models for chest x-rays and CT-scans classification in contrast with a deep CNN model built from scratch. DenseNet121, Resnet-50, Inception v2, Resnet101-V2 and VGG16 are selected and initialized with either random or pre-trained weights to classify augmented images into two classes: Covid and non-Covid. The performance evaluation proves the minuscule impact of training transfer learning models for good quality results, as all CNN models contribute almost equally to the classification and achieve considerable results in terms of precision, accuracy, recall and F1 score.

## 1 Introduction

For almost two years now, the world has been struggling with the coronavirus pandemic that has drastically affected the world economy but, more importantly drag thousands of people to death path. The early diagnosis of infected people is beneficial to avoid the widespread outbreak of the COVID-19 virus. However, this procedure can be affordable to some countries but raises a lot of concerns to others due to the test price and privatization of healthcare systems and facilities. Fortunately, many clinical centers have recently released hundreds of thousands of chest x-ray images to the scientific community, allowing feature pattern recognition of COVID-19 disease [1]. However, the interpretation of radiography images is prone to human error leading to possible false negative diagnosis. Artificial intelligence has been proven to be a game-changer in healthcare through performing an automated data learning and patterns discovery, thus replacing highly delicate tasks performed by radiologists. Convolutions neural network (CNN) are commonly used to analyze medical imagery[2]. Many researches proposed effective CNNs to diagnose infected cases based on chest x-rays and CT-scans. The priority was given to chest x-rays as they are widely available in health centers, however x-rays machines does not show soft tissue injuries or inflammation,therefore chest CT-scans is envisioned as a good alternative to enhance the accuracy of detection. The process of extracting features from x-rays(or ct-scans) and classify them into class diseases requires large-size datasets. The access to Covid-19 medical records can be restricted to the research community, as part of the practice of maintaining the security and confidentiality of patients. In an effort to provide an abundance of data for training and testing deep learning models, some institutions including the US National Library of Medicine have made clinical readings available to the public [3]. Transfer learning has also been extensively used in medical applications for classification purpose. Such pre-trained models is assumed to bypass the need for large-sized datasets by using pre-trained weights from ImageNet-based CNN models [4]. In this paper, the performance of transfer learning models is studied in depth to highlight its feasibility in comparison with deep CNN model trained from scratch.

## 2 Related works

In [5], S.K Venu studied the state-of-the-art deep learning models for pneumonia classification. A weighted average ensemble of Inception, ResNet152V2, DenseNet201, MobileNetV2 and Xception was created, and the evaluation results demonstrated a test accuracy of 98.46%, precision of 98.38%, recall of 99.53%, and f1 score of 98.96%. Arpita Halder and Bimal Datta developed in [6] a deep learning framework that includes the following pre-trained models as a backbone named KarNet : DenseNet201, VGG16, ResNet50V2, and MobileNet. The models were trained with augmented and unaugmented datasets and the results demonstrated good diagnostic ability, with AUC scores of 1.00 and 0.99 for models trained on unaugmented and augmented data sets, respectively. DenseNet201 achieved an accuracy of 97% for the test dataset, followed by ResNet50V2, MobileNet, and VGG16 with accuracies respectively equal to 96%, 95%, and 94%. In [7], authors presented a new model using transfer learning method and InceptionV3 algorithm to classify COVID-19 x-rays into Normal and Pneumonia classes. The experimental results demonstrated the effectiveness of the proposed model which achieved an accuracy of 98%. In [8], Maithra Raghu tackled many points on transfer learning for medical applications. The results proved that transfer learning offers limited performance gains to the classification of chest x-rays. A comparative analysis of the precision, recall, F1 score and accuracy of four different models was presented in [9]. Convolutional Neural Network model, XGBoost, VGG Net model and Support Vector Machine were selected to classify chest x-ray images into three classes:COVID, healthy and pneumonia. The results showed that machine learning models featuring deep learning give a good performance with an average precision, recall, F1-score and accuracy of 95.27%, 94.52%, 94.94%, and 95.81%, respectively.

## 3 Transfer learning for Medical Imagery

Although CNNs had been maintained as a main pillar of computer vision field for decades, training a deep architecture from scratch remains full of challenges. In fact, training a deep Convolutional Neural Network architectures requires considerable computational power and GPU memory, despite this, the batch size might be far from optimal as it becomes smaller when depth gets larger. Moreover, CNN models are data hungry, therefore, providing the required amount of labeled training data for medical use can be very difficult due to the scarcity of data, this is an issue that leaded experts to resort to data augmentation. To resolve the complications of training a deep CNN from scratch, a pre-trained CNN model is fine tuned to transfer knowledge to the new model in hand, thus mitigating the extensive time and resources consumption. Transfer learning is the term referring to the improvement of learning in a new task through the transfer of knowledge from a related task that has already been learned [10]. Transfer learning has been involved in various image analysis projects alongside with deep learning. For instance, Ulysses and .al developed in [11] a Transfer Learning(TL) algorithm in the context of Surface Electrocardiography to make use of the large amount of the hand gesture recordings aggregated from multiple participants, hence offering a large feature pool for the deep learning algorithms to learn from. In [2], researchers from Facebook applied transfer learning for image classification. The TL model consists in training a deep CNN model based on hashtags assigned to gazillions of images on social media and fine-tune the pre-trained model to detect objects or classify images. In biomedical image analysis, collecting the required data to train a deep CNN model for disease pattern recognition is not always possible. Therefore, using pre-trained models can often fill the data-scarcity gap and avoid the need for technical expertise to label biomedical images. The term of homogeneous and heterogeneous transfer learning were presented in detail in [12]. The former solution refers to the deep CNN modeling cases that share the same feature space, whereas the latter imply distinct feature spaces. The concept of relaying knowledge to perform another task inspired many researchers to apply transfer learning to detect patients with Confirmed Coronavirus Disease. In spite of the very promising advantages of applying transfer learning to detect patients with COVID-19, some points remain questionable regarding the convenience of the pre-trained model to medical domain. In fact, transfer learning is built upon a fine-tuned standard ImageNet architecture to perform chest x-rays classification. However, the target task of ImageNet and medical domain have major differences. Covid-19 pneumonia can be seen as whiteness in the lungs on radiography whilst ImageNet dataset mostly include clear and defined object. Moreover, ImageNet is considered as the biggest image dataset containing more than 14 million images of more than 20000 different categories having 27 high-level subcategories containing at least 500 images each, hence, Covid-19 chest x-rays dataset is viewed as a small category subset as it hardly involves hundreds of thousands images. With these considerable differences between the two datatsets, can we approve the feasibility of ImageNet based transfer learning model to even detect low level features in chest x-rays images? In order to find some answers to this question, a set of experiments is performed on ImageNet based transfer learning models. The fine-grained study is given in more details in the following section.

### 3.1 Application and results

#### 3.1.1 Fine-tuning

The performance of the following pre-trained models is thoroughly taken into consideration:Resnet-50, Resnet101-V2, DenseNet121, VGG16 and Inception v3. To re-purpose these pre-trained models for chest X-ray images classification, the original classifier is replaced with a binary classifier. The classifier is fine-tuned according to two strategies: train the entire model and freeze the convolutional base. The first strategy consists in learning the model from scratch by training it in accordance to chest x-rays data-sets, whilst the second strategy keeps the original form of the convolutional bas, and its output is fed to the binary classifier. A third strategy would consider freezing a specific number of layers and training the rest of layers to the new data-set, however the main purpose in this study is not to find the best suited architecture to get a good classifying model rather than to highlight the performance of transfer learning models in their most common used case scenarios. Other factors can be involved in defining the choice of a strategy such as the size of the dataset and the available computational power. The performance of the pre-trained models is compared to the proposed deep CNN model presented in Figure 1. The output shape of each layer and the number of weight parameters are shown in details in Table 1.

**Table 1:** CNN model architecture

| Layer (type) | Output Shape | Param |
| --- | --- | --- |
| SeparableConv2D | (None, 198, 198, 16) | 91 |
| MaxPooling | (None, 99, 99, 16) | 0 |
| SeparableConv2D | (None, 97, 97, 16) | 416 |
| MaxPooling | (None, 48, 48, 16) | 0 |
| BatchNormalization | (None, 48, 48, 16) | 64 |
| SeparableConv2D | (None, 48, 48, 32) | 688 |
| MaxPooling | (None, 23, 23, 32) | 0 |
| BatchNormalization | (None, 23, 23, 32) | 128 |
| SeparableConv2D | (None, 21, 21, 32) | 1344 |
| MaxPooling | (None, 10, 10, 32) | 0 |
| BatchNormalization | (None, 10, 10, 32) | 128 |
| Dropout | (None, 10, 10, 32) | 0 |
| SeparableConv2D | (None, 8, 8, 64) | 64 |
| SeparableConv2D | (None, 8, 8, 64) | 2400 |
| MaxPooling | (None, 4, 4, 64) | 0 |
| SeparableConv2D | (None, 2, 2, 64) | 4736 |
| MaxPooling | (None, 1, 1, 64) | 0 |
| BatchNormalization | (None, 1, 1, 64) | 256 |
| Flatten | (None, 64) | 65000 |
| Dense | (None, 1000) | 0 |
| Dense | (None, 2) | 2002 |

**Fig. 1:**
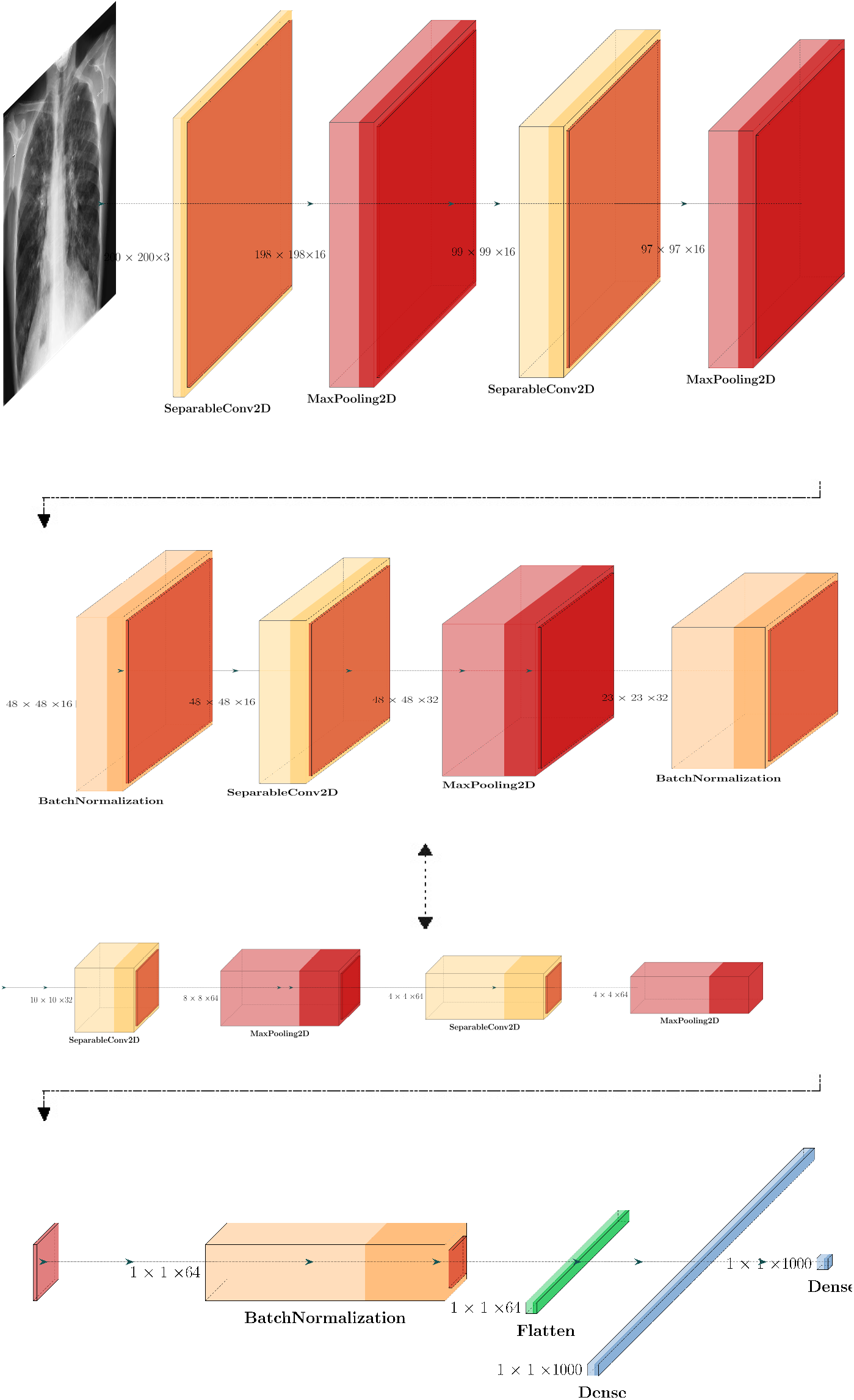
CNN model

#### 3.1.2 Dataset

This performance study uses medical images dataset from two sources. The first COVID-19 dataset consists of Non-COVID and COVID cases of both X-ray and CT images. The dataset is augmented with different augmentation techniques to generate about 17100 X-ray and CT images [13]. The second dataset contains lung CT-scans collected from real patient in Sao Paulo, Brazil[14]. There is a total of 2482 CT scans divided into two classes: COVID(1252)and non-COVID(1230). Both datasets are publically available at https://data.mendeley/com/datasets/8h65ywd2jr/2 and https://www.kaggle.com/plameneduardo/sarscov2-ctscan-dataset. Samples of CT-scans and chest X-ray images for infected and healthy patients are shown in Figure 2 and Figure 3 respectively.

**Fig. 2:**
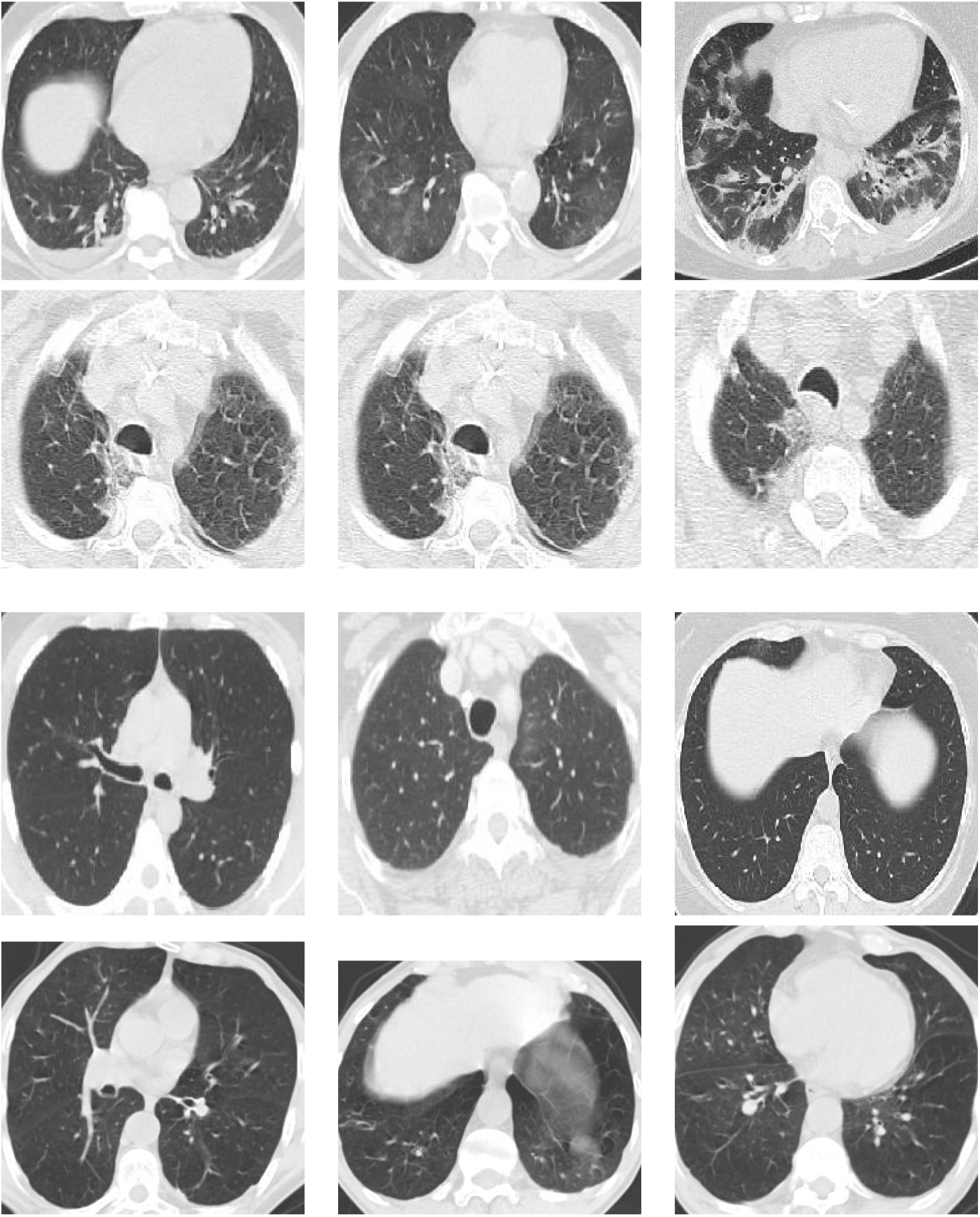
Sample images from CT-scan dataset [14] showing COVID-19-positive (The first two rows) and COVID-negative (The last two rows) scans.

**Fig. 3:**
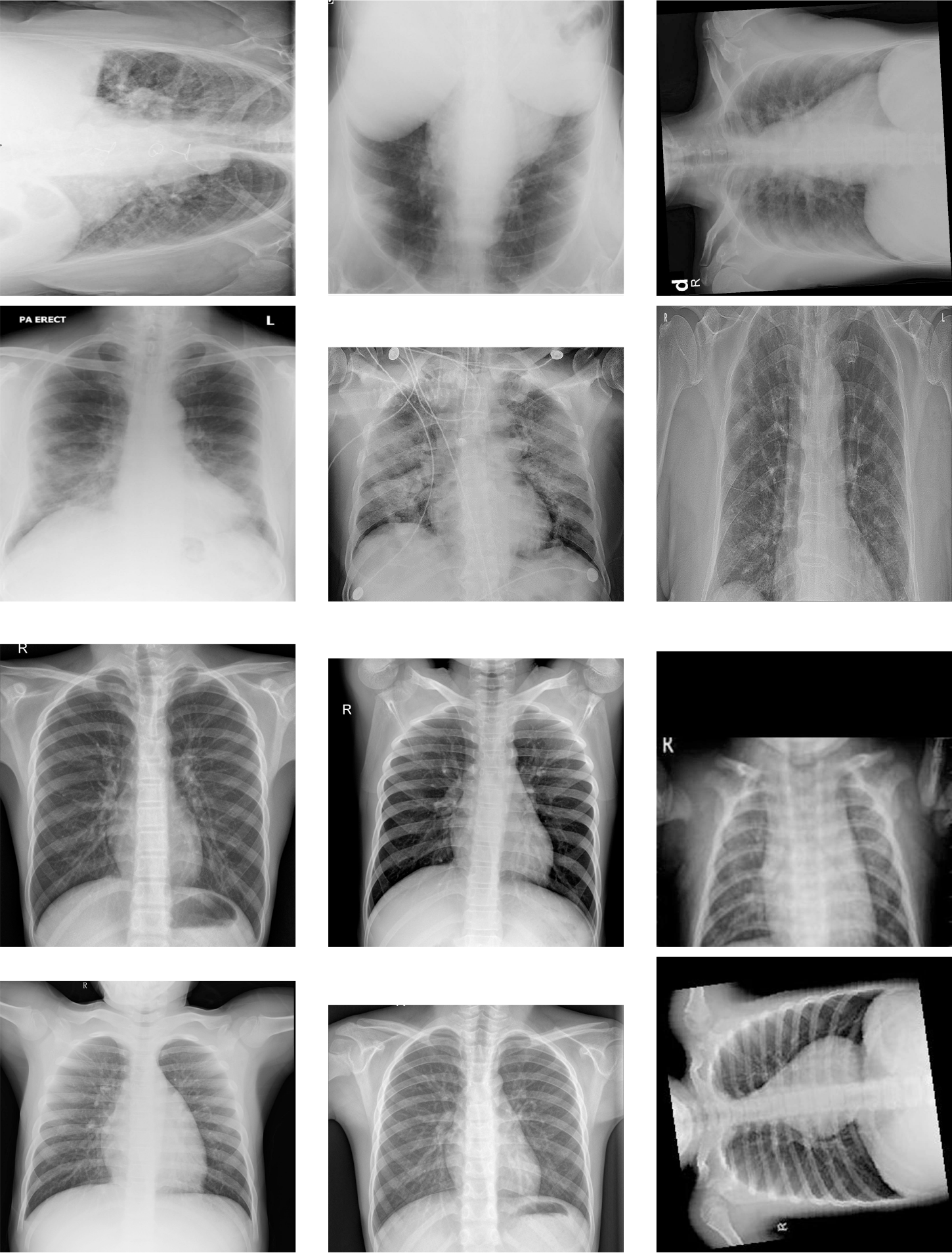
Sample images from chest x-rays dataset [13] showing COVID-19-positive (The first two rows) and COVID-negative (The last two rows) scans

#### 3.1.3 Data pre-processing

The chest x-ray and CT-scan images are augmented and balanced to create a variety of input data and prevent the severely skewed class distribution. Data augmentation extends the training dataset to enhance the performance of the predictive model. One of the major problems encountered while working with large datasets is memory management. In fact, the dataset can be too big to fit into available memory. A solution to this issue consists in loading and augmenting images in batches during the training process. This procedure is called *Data Augmentation*. Data Augmentation allows to create new training instances while maintaining the class type of the base instance. In the present paper, *On The Fly Data Augmentation* method is utilized to augment the training images [15]. The ImageDataGenerator class in Keras [16] is an implementation of this method, where the model is trained on generated instances that vary at each epoch, hence helping the model to better generalize. The resulting number of augmented images is equal to: *the number of training images × the number of epochs*. A rotation augmentation is set to randomly rotate the image between −25 to 25 degrees. A horizontal flip and a zoom augmentation are enabled with a zoom range set to 0.2. The augmented images are resized and normalized between 0 and 1 to fit the requirements for the transfer learning models. The input images are resized to (299 *×* 299 *×* 3) for Inception v3 model and to (224 *×* 224 *×* 3) for Resnet-50, Resnet101-V2, DenseNet121, VGG16 with data format corresponding to “channels-last”. Applying data augmentation on the input images generates a total maximum of 90.000 augmented images. These images are converted to arrays before they are randomly split into training, validation and test sets with the following distribution 80%, 10% and 10%.

#### 3.1.4 Analysis methodology

A deep CNN model performance is compared to transfer learning models with random weights initialization and pre-trained weights. In this study, a new neural network integrates the pre-trained model(with the original classifier removed) and a new binary classifier. The new classifier includes a Flatten layer added after the last pooling layer in each pre-trained model, plus a Dense fully connected layer composed of 1000 neurons. The last output layer predicts the Covid and non-Covid classes. Two usage patterns are considered for performance analysis:Integrated Feature Extractor and Weight Initialization. The first pattern freezes the layers of the pre-trained model during training whilst the second initializes the weights randomly and the model is trained from scratch.

#### 3.1.5 Performance Metrics

Various evaluation metrics are utilized to assess the performance of the deep learning models. All the following metrics used in this study are based on the confusion matrix shown in Figure 4: recall, precision, F1 score, micro recall, micro precision, micro F1 score and accuracy.

**Fig. 4:**
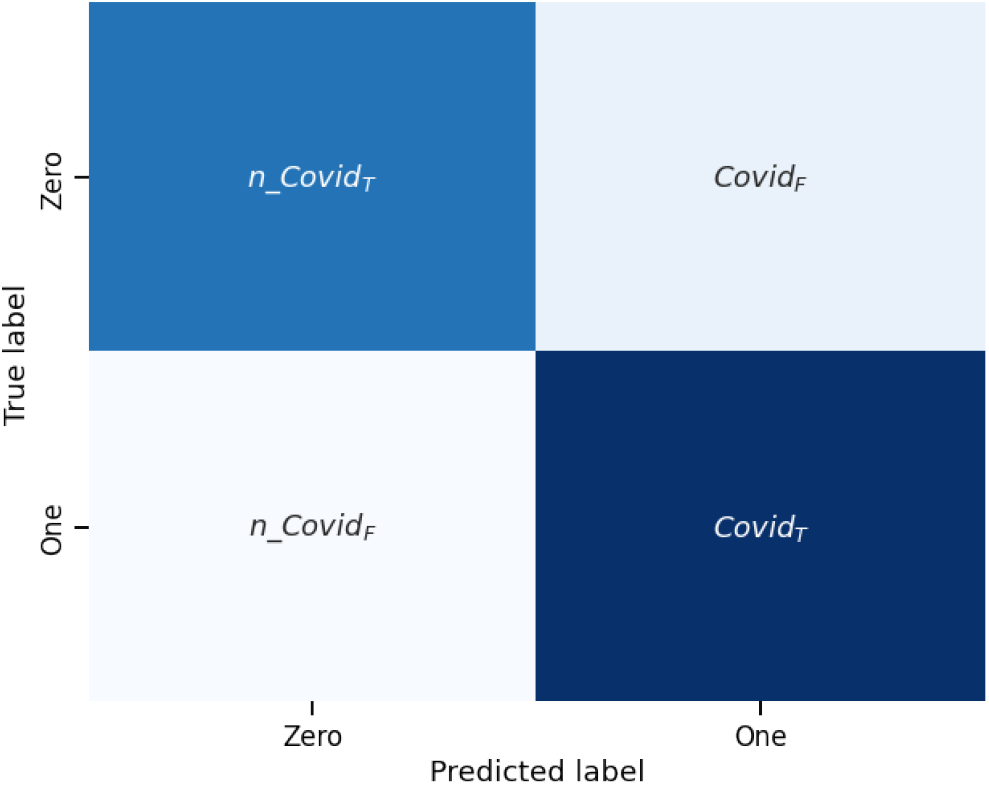
Confusion Matrix for *Covid*19 coronavirus disease detection

where :

- *n - Covid*_*T*_ :True Positive covid cases.
- *Covid*_*T*_ :False Negative covid cases.
- *n - Covid*_*F*_ :False Positive covid cases.
- *Covid*_*F*_ :True negative covid cases.

Recall is a metric used to define the proportion of actual covid-19 positive cases that were predicted correctly. The formula is depicted below:

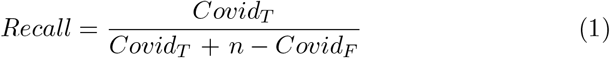

Precision is a metric used to define What proportion of positive predictions that were actually correct. Precision formula is defined as follows:

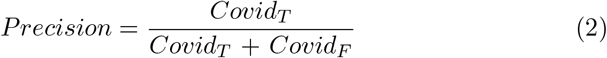

F1-score metric assess the quality of binary classification problems. It is often used to select a model based on a balance between recall and precision. The F1-score metric is defined as:

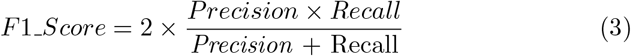

Micro-recall or micro-averaged recall measures the recall of the aggregated contributions of Covid and non-Covid classes. The Micro-recall equals to:

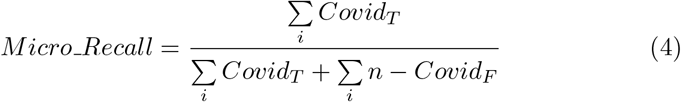

Micro-precision or micro-averaged precision measures the precision of the aggregated contributions of all Covid and non-Covid classes. The formula is depicted below:

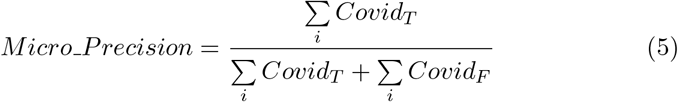

Micro F1-score assess the quality of multi-label binary problems. It measures the F1-score of the aggregated contributions of Covid and non-Covid classes. Micro F1-score is defined as:

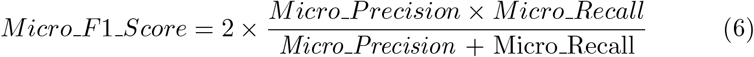

The accuracy metric measures how often the model predicts correctly. The accuracy formula is defined as:

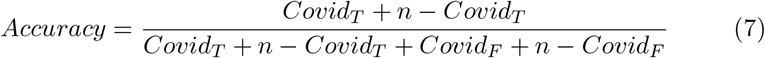

The experiments are conducted on a Virtual machine having 13GB RAM, 33GB HDD and 2-core Xeon 2.2GHz. All models are trained with approximately 90.000 augmented images divided into training, validation and test sets. The batch size is set to 32 and the loss function is optimized with Adam optimizer. The learning rate is set to 1 *× e*^*-*5^ for VGG16 model to converge quickly to the optimal values and to 1 *× e*^*-*3^ for the rest of the models. The callback function is utilized to access the internal state of the model and stop training when the validation accuracy reaches 0.89 and the validation loss is below 0.3 to prevent an over-fitted model. All models are implemented using Keras framework built on top of TensorFlow 2. The performance study considers both chest x-rays images and CT-scans. The experimental results are presented in Figure 5 and Figure 6.

**Fig. 5:**
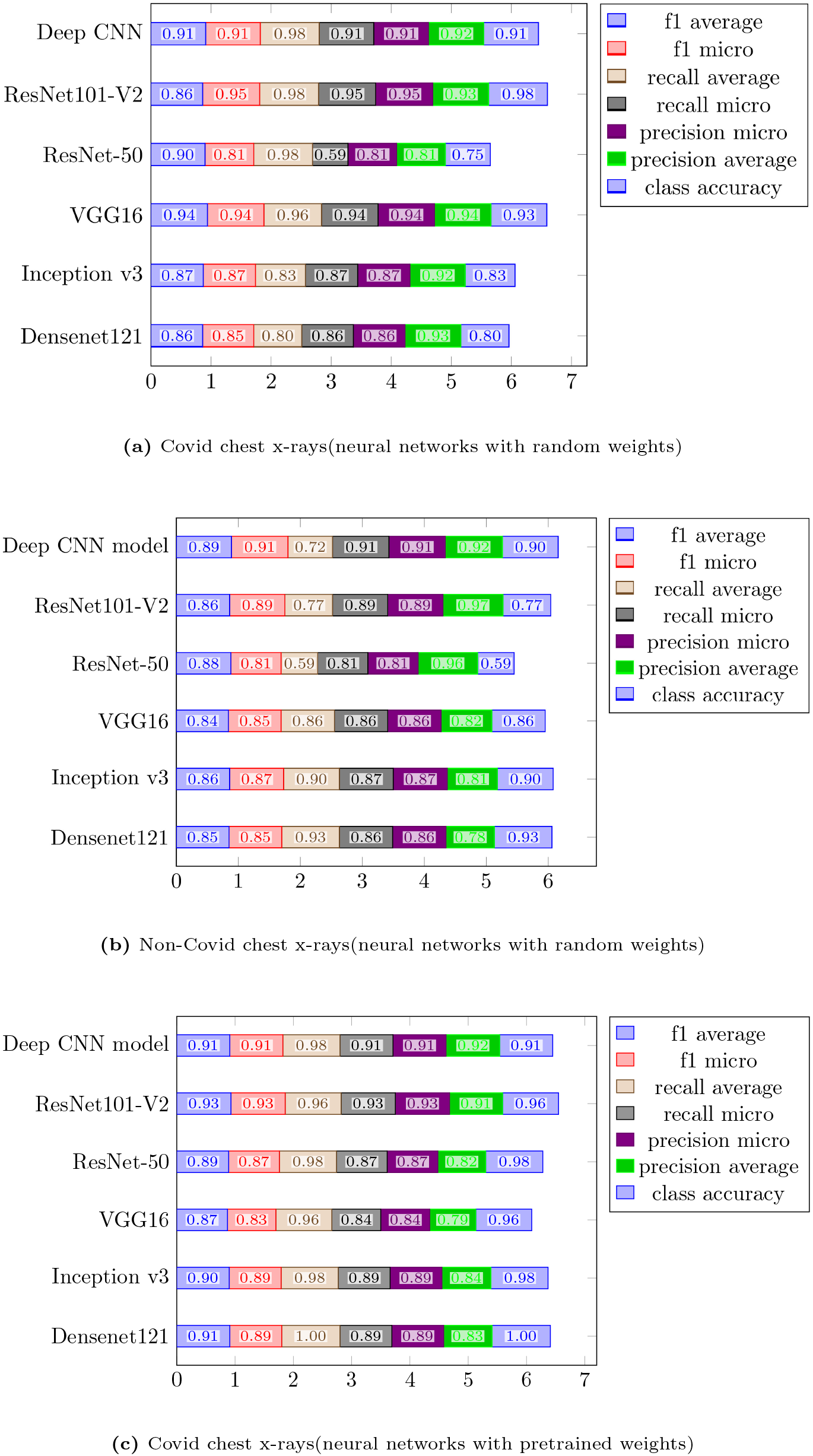
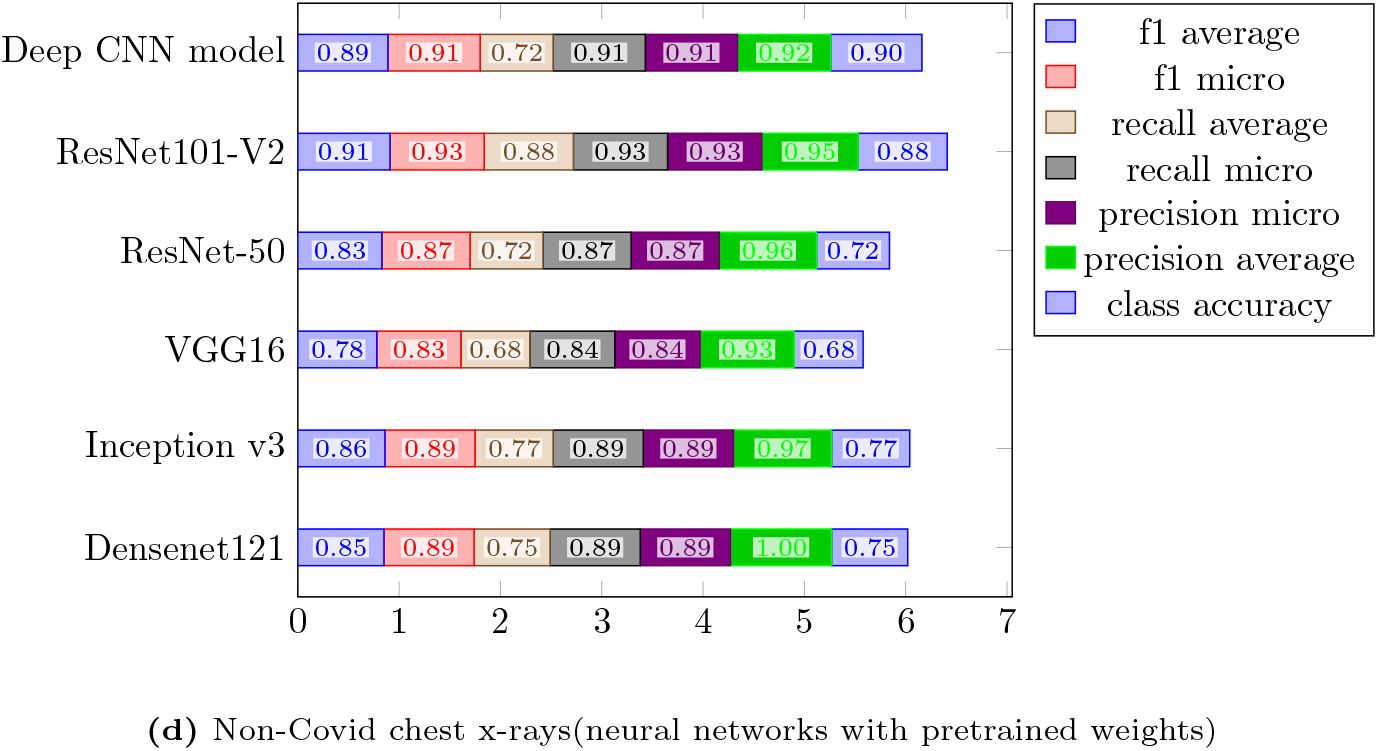
Experimental results of deep learning models with chest x-rays images

**Fig. 6:**
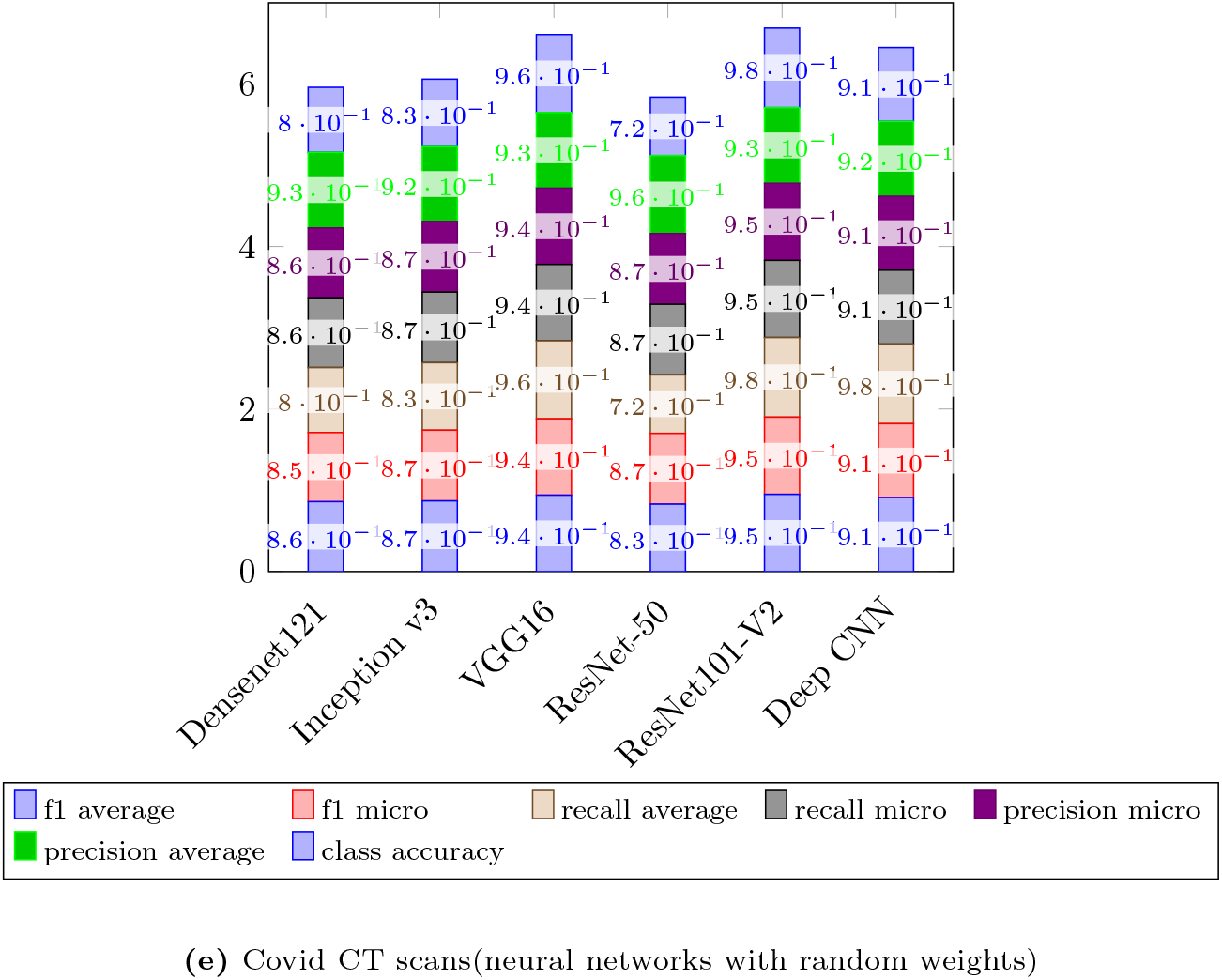
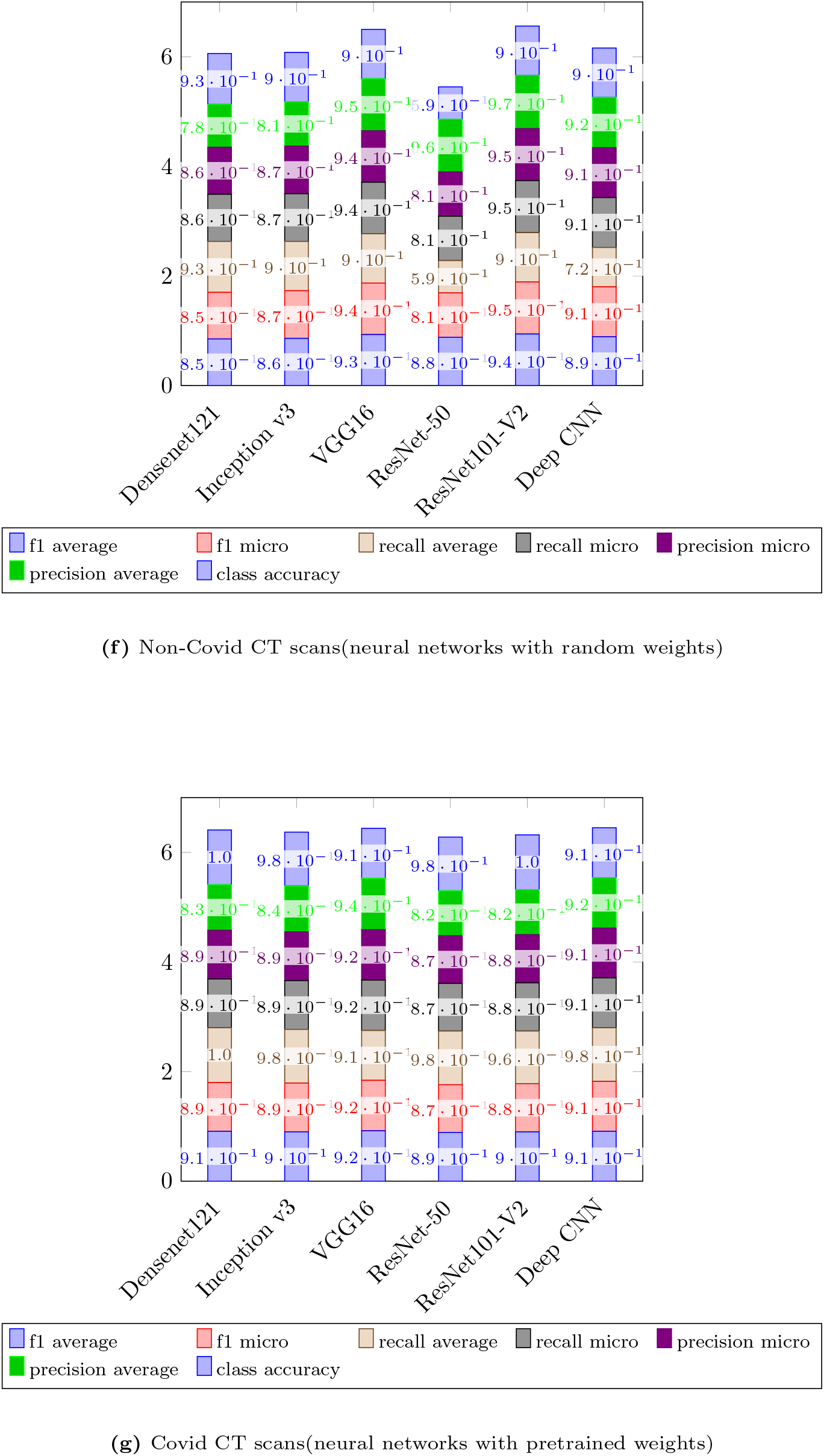
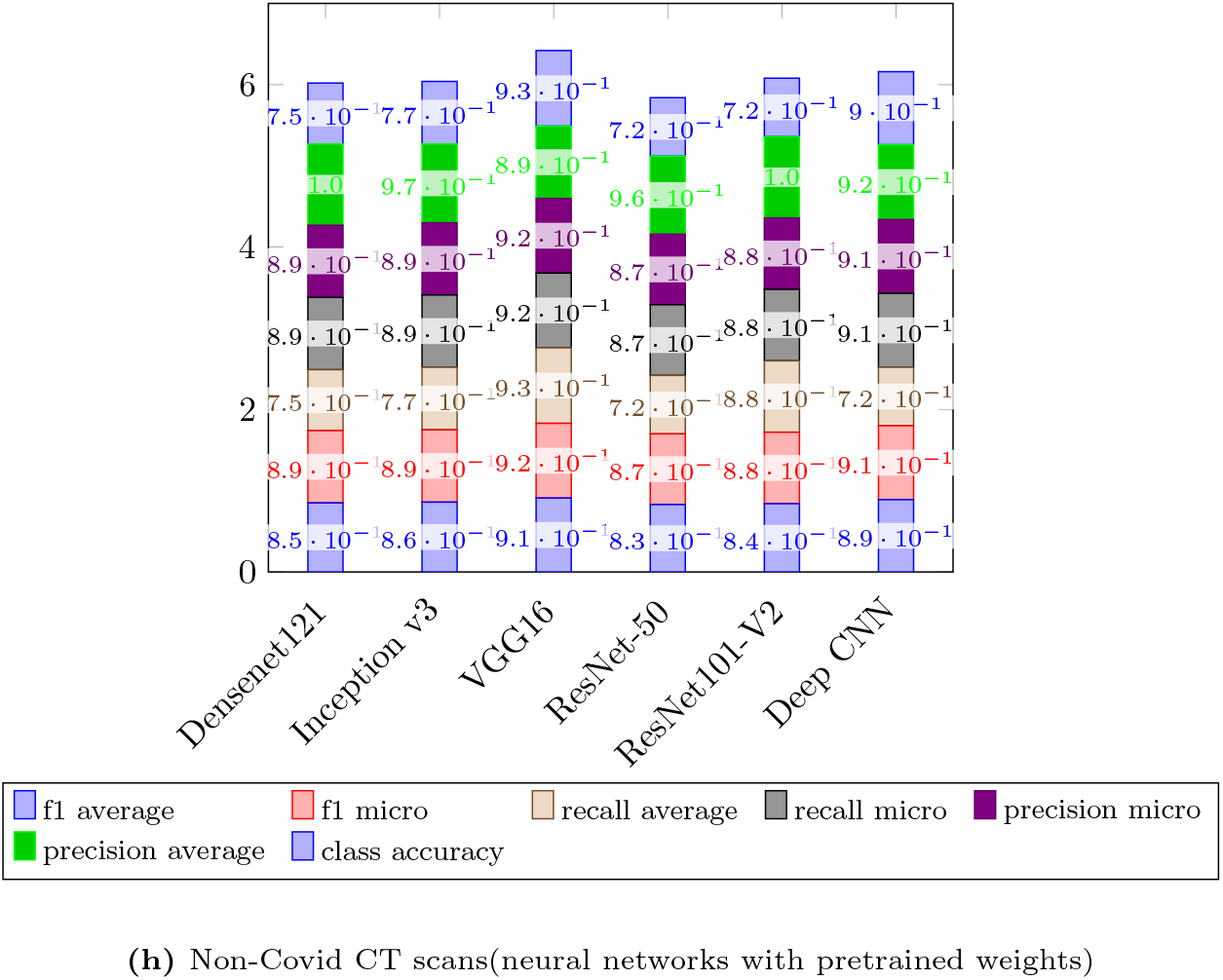
Experimental results of deep learning models with CT scans

#### 3.1.6 Result and Discussion

The results from Figure 5 and Figure 6 indicate that all the trained models have successfully classified Covid-19 patients from non-Covid-19 patients on the test set, with a slight differentiation in the evaluation metrics values. The maximum accuracy 98% is achieved by Densenet121 and ResNet-50 model that were initialized to pre-tained and random weights. Densenet121 model has the highest recall with 100% of actual covid-19 positives cases predicted correctly, whilst the least recall performance was given by the deep CNN model with only 72% of cases predicted correctly as healthy patients. ResNet-50 scored 96% for the proportion of positive predictions that were actually infected with the disease versus 95% for VGG16. In contrast, as can be shown in Figure 7, the number of epochs needed to train the deep CNN model is equal to 8 versus 405 epochs for VGG16 with random initialized weights, and 5 epochs for pre-trained Densenet121. All the trained models achieved approximately similar performance when classifying healthy patients and patients with covid-19. The overall variance of the evaluation metrics considering all models ranges between 0.02 and 0.28. This difference in contribution is believed to be small and leaves the choice open to either use a transfer learning model or train a deep CNN model from scratch for chest x-rays and CT-scans classification. Nevertheless, the proposed deep CNN model outperformed the transfer learning models in terms of epochs number as an increasing number of epochs tends to over-fit the CNN model which leads to a poor performance on new images dataset.

**Fig. 7:**
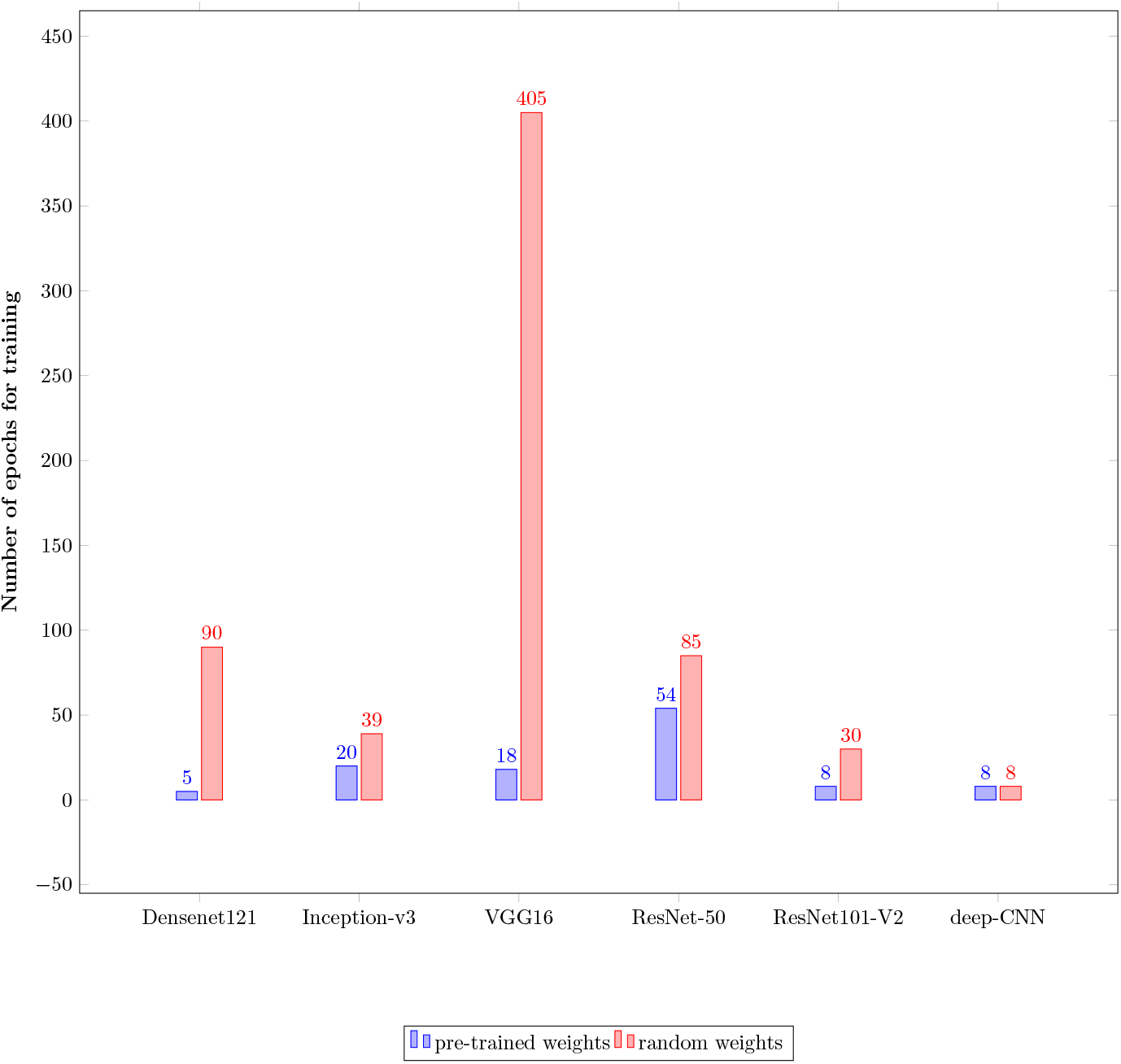
Total number of epochs

## 4 Conclusion

In this paper, the feasibility of transfer learning for medical images classification has been investigated. The results demonstrated the limited contribution of transfer learning models to the classifications of chest x-rays and CT-scans into Covid and non-Covid classes. A deep CNN model trained from scratch is proven to show a performance similar to transfer-based models with less number of epochs and small architecture. The healthcare data raises the following two major challenges when dealing with deep learning models: a limited medical dataset and a different feature context that mainly includes natural image features from ImageNet. Data augmentation techniques were applied to augment images with different sub-transformation including zooming, translation, shearing and rotation. It is worth mentioning that transfer learning models do not always bring positive results when the source and the target tasks belong to different domains. A good practice is to use pre-trained model over ImageNet data-sets as medical images are not included in ImageNet, or train a deep CNN model from scratch over medical images to build transfer learning models for medical applications.

## Data Availability

https://data.mendeley.com/datasets/8h65ywd2jr/2
https://www.kaggle.com/plameneduardo/sarscov2-ctscan-dataset

## Compliance with Ethical Standards

- Conflict of interest:Author Maryam El Azhari declares that he/she has no conflict of interest.
- Ethical approval: This article does not contain any studies with human participants performed by any of the authors.

## Notes

### Competing Interest Statement

The authors have declared no competing interest.

### Funding Statement

This study did not receive any funding

### Author Declarations

https://data.mendeley.com/datasets/8h65ywd2jr/2 https://www.kaggle.com/plameneduardo/sarscov2-ctscan-dataset

